# Identifying Neuroanatomical and Behavioral Features for Autism Spectrum Disorder Diagnosis in Children using Machine Learning

**DOI:** 10.1101/2020.11.09.20227843

**Authors:** Yu Han, Donna M. Rizzo, John P. Hanley, Emily L. Coderre, Patricia A. Prelock

## Abstract

Autism spectrum disorder (ASD) is a neurodevelopmental disorder that can cause significant social, communication, and behavioral challenges. Diagnosis of ASD is complicated and there is an urgent need to identify ASD-associated biomarkers and features to help automate diagnostics and develop predictive ASD models. The present study adopts a novel evolutionary algorithm, the conjunctive clause evolutionary algorithm (CCEA), to select features most significant for distinguishing individuals with and without ASD, and is able to accommodate datasets having a small number of samples with a large number of feature measurements. The dataset is unique and comprises both behavioral and neuroimaging measurements from a total of 28 children from 7 to 14 years old. Potential biomarker candidates identified include brain volume, area, cortical thickness, and mean curvature in specific regions around the cingulate cortex, frontal cortex, and temporal-parietal junction, as well as behavioral features associated with theory of mind. A separate machine learning classifier (i.e., k-nearest neighbors algorithm) was used to validate the CCEA feature selection and for ASD prediction. Study findings demonstrate how machine learning tools might help move the needle on improving diagnostic and predictive models of ASD.

## Introduction

Autism Spectrum Disorder (ASD) is a neurodevelopmental disorder that can cause significant social, communication, and behavioral challenges. According to the most recent report from the Center for Disease Control and Prevention (CDC), the number of children in the U.S. diagnosed with ASD are about 1 in every 54 [80]. While genetic and environmental factors have been linked to the development of ASD, at present there is no identified cause or cure of ASD.

ASD is characterized by impairments in social interaction and the presence of restricted and repetitive behaviors, interests, or activities [1–4]. At the core of social impairment is often a deficit in theory of mind (ToM), the ability to recognize and understand the thoughts, feelings and perspectives of others. Some symptoms of ASD are not evident until age two or later. In fact, a child may appear to be developing according to typical milestones until the age of two and then stop learning new skills and may even lose skills [5, 6]. Currently, the diagnosis of autism is based on behavioral symptoms alone, often assessed through the Autism Diagnostic Observation Schedule-Second edition (ADOS-2) and the Autism Diagnostic Interview-revised (ADI-R) [7, 8]. A typical diagnostic appointment consists of evaluations lasting several hours at a designated clinical office. The rigorous and time-consuming nature of ASD diagnostic examinations often leads to a demand that exceeds the capacity to see patients. As a result, many diagnostic centers have expanding wait lists for appointments. This bottleneck can translate to delays in diagnosis of 13 months and longer [5, 9–15]. It is also believed that a substantial number of individuals on the spectrum remain undetected [82]. With growing awareness of ASD, there is a high demand for a faster and more automated ASD diagnostic approach that might allow for more efficient diagnosis and early identification of high-risk populations [16].

Building an automated diagnostic and predictive model of ASD is timely as many studies have adopted machine learning approaches to identify significant biomarkers that include both behavioral and biological features. For instance, Duda and colleagues (2016) applied machine learning to distinguish ASD from attention deficit hyperactivity disorder (ADHD) using the Social Responsiveness Scale for children between 5 to 13 years old [17]. Bone et al. (2015) trained their models to diagnose children with ASD versus neurotypical (NT) children using the same Social Responsiveness Scale and the ADI-R score for children between 5 to 17 years old [18]. Other studies aggregated items from the ADOS and scores from the Autism Quotient (AQ) to accurately classify an ASD group [19]. Although behavioral outcome measures are commonly used for ASD diagnosis and assessment with solid reliability and validity, there are some limitations when using them to classify participants. Specifically, behavioral outcome measures are typically prone to performance demands, as well as environmental and personal factors that could influence responses on any given day. Consequently, the ability to identify more consistent ASD markers using neuroimaging measures to supplement behavioral measures becomes important.

As a result of the wide range and subjective nature of behavioral measures used in diagnosing ASD, many studies are exploring brain-based biological markers (e.g., measurable via magnetic resonance imaging (MRI)) to identify a common etiology across individuals with ASD. Currently, these less subjective markers are attractive not only for diagnostic purposes, but as possible targets for interventions [20]. Independent structural MRI studies have found differences in whole brain volume and the developmental trajectories between individuals with and without ASD [21–29]. Other structural brain abnormalities associated with ASD include cortical folding signatures that appear in the following regions of the brain: temporal-parietal junction, anterior insula, posterior cingulate, lateral and medial prefrontal cortices, corpus callosum, intraparietal sulcus, and occipital cortex [21–29]. Evidence also shows that an accelerated expansion of cortical surface area, but not cortical thickness, causes an early overgrowth of the brain in children with ASD [30], while other studies suggest that individuals with ASD tend to have thinner cortices and reduced surface area as an effect of aging [31]. With these clear brain differences among those with and without ASD, it is informative and critical to look for brain-based ASD biomarkers.

Machine learning (ML) has been introduced to the neuroimaging field to identify the atypical brain regions in individuals with ASD. The support vector machine (SVM) is an algorithm that avoids overfitting and is known for high classification accuracy without requiring large sample sizes. It has been used to classify ASD from NT participants using extracted features from functional connectivity metrics and grey matter volume [32–35]. Other ASD applications of ML classifiers include deep neural networks [37] and the random forest (RF) algorithm; the latter uses random ensembles of independently grown decision trees [38]. Although these methods have demonstrated high accuracy for classifying ASD, to our knowledge most of them have not been used to identify input variables most closely associated with ASD (i.e., feature selection). In addition, the majority of studies use data from the Autism Brain Imaging Data Exchange (ABIDE) dataset, which includes 1112 existing resting-state functional MRI (rs-fMRI) datasets with corresponding structural MRI and phenotypic information from 539 individuals with ASD and 573 age-matched NT controls between the ages of 7 and 64 collected from 24 international brain imaging laboratories [39]. For example, Guo et al. (2017) selected features associated with ASD using a deep neural network from brain resting-state functional connectivity patterns using the ABIDE dataset [83]. However, findings using datasets from various sites have several limitations.

Specifically, classification across a heterogeneous population as is true in autism is challenging [40, 41], particularly when neuroimaging data are pooled from multiple acquisition sites such as the ABIDE dataset, which has considerable variation in demographic and phenotypic profiles. Data variance introduced via scanner hardware, imaging protocols, operator characteristics, regional demographics, and other site-specific acquisition factors can also affect the classification performance. It is often difficult to collect neuroimaging data from individuals with ASD given the scanner noises and high expectations for individuals to remain still. In fact, most individual site datasets have small sample sizes that can lead to overfitting and classification inaccuracies using traditional ML algorithms. Moreover, while the ultimate goal for ML-based diagnostic classification in neuroimaging is to identify discriminative features that provide insight into atypical structure and connectivity patterns in the affected population [42], many of the ML algorithms applied to ASD were designed to classify large amounts of data (e.g., ABIDE) rather than optimize the selection of input features.

ASD drivers or markers are likely the result of a complex interaction of factors with no single factor (i.e., main effect or univariate model) driving the system. As such, traditional statistical tools (e.g., logistic regression) that search for univariate drivers of ASD are unlikely to find consistent patterns. Thus, ML techniques that explore large search spaces for multivariate interactions are both needed and becoming popular in helping to elucidate the complex interactions in systems such as ASD. Our study employs one such ML tool: an evolutionary algorithm [44] called the conjunctive clause evolutionary algorithm (CCEA) [36]. The CCEA was specifically designed to efficiently explore large search spaces for complex interactions between features and some associated nominal outcome (e.g., ASD or NT). In addition, the CCEA has built-in tools to prevent overfitting to produce easily interpretable parsimonious models.

This study examines the utility of using the CCEA for feature selection in ASD, particularly to address traditional statistical challenges associated with datasets having small sample sizes and a large number of feature measurements. Additionally, the selected features are validated and used for diagnostic classification by applying a separate and more traditional ML classifier (i.e., the k-nearest neighbors (KNN) algorithm). The dataset in the present study has a relatively large number of features, consisting of both behavioral and neuroimaging measurements. Although some behavioral features may seem less objective compared to an assessment of neuroanatomical features, they are often easier to obtain and more pragmatic for children with ASD especially if they have strong psychometric properties. Clinicians seldom place a child with ASD in a scanner to obtain neuroanatomical information before conducting behavioral assessment for the purpose of diagnosis and treatment. The combination of behavioral measures with neuroanatomical information supports the value of making brain-behavior connections that will advance our understanding of ASD. In the present study, the behavioral measurements include scores of language ability, intellectual ability, and ToM. The neuroimaging measurements include brain volume, brain surface area, cortical thickness, and cortical curvature extracted from MRI whole-brain T1-weighted scans. These features were collected from a total of 28 children ages 7 to 14, of which 9 children had been diagnosed with ASD. Only a subset of these 28 children were used for feature selection in the CCEA (7 children with ASD and 14 NT children), as another subset (2 children with ASD and 5 NT children) were enrolled at a later time (i.e., after the CCEA was trained). While this later cohort was not included in the CCEA feature selection analysis, it was included in the subsequent validation and predictive k-nearest neighbors (KNN) modeling.

Using the CCEA, we aim to identify discriminative biomarkers and behavioral features to help develop an automated diagnostic and predictive system for ASD. We believe this is a pioneering study for:

- Selecting discriminative biomarkers among children from 7 to 14 years old and classifying ASD.
- Including both behavioral and neuroimaging measurements in the feature selection model for better prediction and understanding of ASD.
- Identifying models (sets of features) that most strongly correlate to children with ASD given a dataset with a relatively small sample size (i.e., N=28) and a large number of features (i.e., 247 neuroimaging features and 13 behavioral features).
- Developing a predictive ML model using input features selected by the CCEA.

## Materials and Methods

### Participants

A total of 9 children with ASD (1 female) and 19 NT children (7 female), ages 7-14, were enrolled in the study. In addition to the behavioral assessments described below, the ASD group also completed the ADOS-2 and the Social Communication Questionnaire-Lifetime version (SCQ) [45] to confirm their ASD diagnosis. Although diagnosis of ASD is typically done at an early age, the characteristics of ASD are long-term, and classification with additional neurobiological information at any age recognizes the potential for brain-behavior comparisons with NT populations. Potential changes behaviorally and neurobiologically at any age may also inform types, duration, and intensity of intervention that may influence these changes. Therefore, although we tested older children, this study informs an increased understanding of likely biomarkers of ASD.

### Ethical Statement

The present study was approved by the University of Vermont (UVM) Research Protection Office and IRB committee. The specific staff member who approved the study protocol, 19-0005, was Karen Crain. Consent forms were obtained from each participant’s legal guardian and assent forms were obtained from each participant.

### Behavioral Measurements

All children participated in 2-3 hours of baseline behavioral assessments that included the Comprehensive Assessment of Spoken Language (CASL) [81], the Universal Nonverbal Intelligence Test-2 (UNIT-2) [46, 47], the Theory of Mind Task Battery (ToMTB) [48] and the Theory of Mind Inventory-2 (ToMI-2) [49, 50]. Measures of language and cognition are typical in the assessment of ASD, as the Diagnostic and Statistical Manual of Mental Disorders (DSM-5) requires an assessment of language and intellectual functioning beyond the diagnosis of ASD.

The CASL is an orally administered research-based assessment consisting of 15 subtests measuring language for individuals ranging from 3 to 21 years of age. For the present study, only those basic subsets that establish the CASL language core are used: Antonyms, Sentence Completion, Syntax Construction, Paragraph Comprehension, and Pragmatic Judgment. Reliability for the core subtests ranges from 0.80 to 0.90 with composite scores ranging from 0.92 to 0.96. Test-retest reliability for individual subtests range from 0.65 to 0.95 with core composite coefficients from 0.92 to 0.93. The UNIT-2 is a multidimensional assessment of intelligence for individuals with speech, language, or hearing impairments. It consists of nonverbal tasks that test symbolic memory, non-symbolic quantity, analogic reasoning, spatial memory, numerical series, and cube design. Reliability studies include internal consistency, test-retest, and scorer reliability. Subtest coefficients range from 0.88 to 0.96 and 0.93 to 0.98 for the composites. Content, criterion and construct validity were largely correlated with other cognitive measures and demonstrated the value of the UNIT-2 for the assessment of diverse groups of children.

ToM is a core deficit in ASD that is often used to explain the social impairments of the disorder. ToM is the ability to reason about the thoughts and feelings of self and others, including the ability to predict what others will do or how they will feel in a given situation on the basis of their inferred beliefs [51, 52]. Scores from both the ToMTB and ToMI-2 were included to provide representative measures of a child’s social cognition level. The ToMTB and ToMI-2 are two norm-referenced tools and behavioral tasks used as outcome measures to assess ToM [51, 52]. Scores from both the ToMTB and ToMI-2 provide valid representations of a child’s social cognition level. The ToMI-2 is a parent-informant measure of a child’s functional level of ToM. Each of the 60 items assesses a particular ToM dimension using items that range from simple content to those that evaluate more complex skills. Each item is rated on a 20-unit continuous scale anchored by “Definitely Not” and “Definitely.” Respondents indicate their response with a vertical hash mark at the point on the scale that best reflects their attitude. Item, subscale, and composite scores range from 0-20 with higher values reflecting greater parental confidence that the child possesses a particular ToM skill. The ToMI-2 is designed to be a socially and ecologically valid index of ToM as it occurs in everyday social interactions. It has demonstrated excellent test-retest reliability, internal consistency, and criterion-related validity for both NT and ASD children as well as contrasting-group validity and statistical evidence of construct validity (i.e., factor analysis). The ToMTB directly assesses a child’s understanding of a series of scenarios tapping ToM. It consists of 15 test questions within nine tasks, arranged in ascending difficulty. Tasks are presented as short vignettes that appear in a story-book format. Each page has color illustrations and accompanying text. For all tasks, children are presented with one correct response option and three plausible distractors. Memory control questions are included that must be passed for credit on the test questions. The ToMTB demonstrates excellent internal consistency and inter task agreement (*α* = 0.91 at T1 and *α* =0.94 at T2) and strong construct validity (*r* = 0.66, *p* < .01) [48, 50, 53].

In selecting potential features for the CCEA, we included 13 behavioral features. These included the total score of the CASL, full scale score of the UNIT-2, abbreviated score of the UNIT-2, total score of the ToMTB, total composite mean of the ToMI-2 (i.e., assessing overall ToM ability), early subscale mean of the ToMI-2 (i.e., assessing early-developing ToM abilities such as regulating desire-based emotion and recognition of happiness and sadness), basic subscale mean of the ToMI-2 (i.e., assessing basic ToM ability such as recognition of surprise), advanced subscale mean of the ToMI-2 (i.e., assessing advanced ToM ability such as recognition of embarrassment). Drawn from a larger study about emotion recognition and ToM, we also included scores from single ToMI-2 items assessing recognition of simple emotions such as happiness and sadness, as well as more complex emotions such as surprise and embarrassment, which ASD children often find difficult to recognize and process [54–56]. Table 1 provides an overview of scores on the 13 behavioral measures. Results from independent sample *t*-tests found that NT participants scored significantly higher (*p<*0.05) than ASD participants on the CASL, UNIT-2 full scale, ToMTB, ToMI-2 total, ToMI-2 early subscale, ToMI-2 basic subscale, ToMI-2 advanced subscale, as well as ToMI-2 single items of surprise, embarrassment and desire-based emotion. In its psychometric development, the ToMI and ToMTB were tested against children with and without autism with scores on both measures, discriminating those children with a diagnosis of ASD from those without such a diagnosis. These measures of ToM also showed a delay in development of early developing and basic ToM skills and a failure to achieve more advanced ToM skills [48,49,50].

**Table 1.** Participant Behavioral Assessments Scores: NT vs. ASD.

|  |  | NT Group<br>(n=19) | ASD Group<br>(n=9) | Group<br>Difference |
| --- | --- | --- | --- | --- |
| *Age* | | 10.2 (7-14) | 11 (8-13) | $p = 0.36$ |
| *CASL* | | 113 (89-132) | 78.8 (40-121) | $p < 0.001^{**}$ |
| *UNIT-2 Full Scale* | | 111.9 (100-135) | 96.2 (56-127) | $p < 0.05^*$ |
| *Unit-2 Abbreviation* | | 110.9 (91-124) | 102 (67-121) | $p=0.16$ |
| *ToMTB* | | 13.3 (11-15) | 11.1 (5-14) | $p < 0.05^*$ |
| *ToMI-2* | *Total* | 17.2 (11.5-19.3) | 12.4 (9.1-17.6) | $p < 0.001^{**}$ |
| | *Early Subscale* | 17.7 (13.8-20) | 14.6 (11.6-17.9) | $p < 0.05^*$ |
| | *Basic Subscale* | 18 (13.2-19.7) | 14 (8.5-19.2) | $p < 0.001^{**}$ |
| | *Advanced Subscale* | 16 (8.3-18.9) | 9.4 (6-16.4) | $p < 0.001^{**}$ |
| | *Happy* | 18.5 (14.1-20) | 17.3 (12.5-20) | $p = 0.24$ |
| | *Sad* | 17.5 (7.1-20) | 16.9 (13.1-20) | $p = 0.62$ |
| | *Surprise* | 17.8 (12.2-20) | 15.3 (10.3-20) | $p < 0.05^*$ |
| | *Embarrassment* | 15.3 (5.8-20) | 10.3 (4.8-20) | $p < 0.05^*$ |
| | *Desire-Based* | 18.9 (15-20) | 16.7 (14.6-20) | $p < 0.05^*$ |

### MRI Acquisition and Preprocessing

All data were acquired using the MRI Center for Biomedical Imaging 3T Philips Achieva dStream scanner and 32-channel head coil at the University of Vermont (UVM). Parameters for T1 acquisition were TR 6.4s, TE 2.9s, flip angle 8 degree, 1mm isotropic imaging resolution with a 256 × 240 *mm2* field of view and 225 slices. Participants watched three videos at home before coming to the MRI center. The first was a cartoon video explaining what an MRI is, and what one might experience while lying in an MRI scanner [58]. The second video, recorded at the UVM MRI mock scanner room, helped visualize the real setting and procedures a child would experience. The third video explained the procedures of wearing earplugs. All participants practiced laying still and became familiar with the scanner noise in the mock scanner room. The T1 structural scan was preprocessed using the Human Connectome Project (HCP) minimal preprocessing pipelines, including spatial artifact/distortion removal, surface generation, cross-modal registration, and alignment to standard space. These pipelines are specially designed to capitalize on the high-quality data offered by the HCP. The final standard space makes use of a recently introduced CIFTI file format and the associated grayordinates spatial coordinate system. This allows for combined cortical surface and subcortical volume analyses while reducing the storage and processing requirements for high spatial and temporal resolution data [60]. Brain anatomical features were extracted using FreeSurfer aparcstats2tabl script [59], which reads brain parcellation statistics directly from T1 scan and exports them to excel sheets. These extracted anatomical features include volume, cortical thickness, mean curvature, and area of all ROIs for each subject. These ROIs were defined using the automatic segmentation procedures that assign one of 37 labels to each brain voxel, including left and right caudate, putamen, pallidum, thalamus, lateral ventricles, hippocampus, and amygdala [61]. There were 276 brain features included in total.

### Conjunctive Clause Evolutionary Algorithm

We used an evolutionary algorithm to identify the features associated with ASD. The CCEA is a machine learning tool that searches for both the combinations of features associated with a given category (e.g., ASD) as well as their corresponding range of feature values [36]. The CCEA can find feature interactions even in the absence of main-effects, and can, therefore, identify feature combinations that would be difficult to discover using traditional statistics. The CCEA selects for the best conjunctive clauses (CCs) of the form:

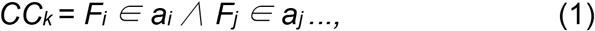

where *F*_*i*_ represents a risk factor i whose value lies in the range *a*_*i*_; and the symbol *∧* represents a conjunction (i.e., logical AND). One benefit of the CCEA is that it produces parsimonious models that are correlated with a select category (e.g., ASD). The models generated by the evolutionary algorithm can be described by their order or total number of features in the conjunctive clause. One example of a parsimonious second order conjunctive clause is: a person with a right hemisphere isthmus cingulate volume of 3,300 – 4,100 *mm*^3^ AND a right hemisphere posterior cingulate volume of 4,100 – 6,200 *mm*^3^ is more likely to have ASD than someone who does not meet these criteria. The CCEA was able to generate models that ranged from 1st to 5th-order in the present study. Since we are only interested in the most parsimonious (i.e., lowest order models) to draw meaningful conclusions and to avoid the overfitting that often occurs with higher-order models, we focused on second-order models (i.e., those having only two features) with the highest fitness (PMF) in which all of the features identified were brain anatomical features. Because of our desire to explore the predictive capability of the behavioral features, we also expanded our analysis to include third-order models (i.e., model combinations with three features) in which behavioral features were included.

The fitness of each conjunctive clause (CC) is evaluated using the hypergeometric probability mass function (PMF), and only the most-fit conjunctive clauses are saved. The hypergeometric PMF is not a *p*-value and thus, is not constrained by issues associated with what threshold is “significant” [62–64]. To prevent overfitting, the CCEA performs feature sensitivity on each conjunctive clause to ensure each feature contributes to the overall fitness. The sensitivity of each feature is calculated by taking the difference between the conjunctive clause fitness and the fitness when that feature is removed. Thus, a feature’s sensitivity may be viewed as the amount of fitness that it contributes to the conjunctive clause. Positive predictive value (PPV) is the number of true positives divided by the sum of true and false positives; and class coverage is the number of true positives divided by the sum of true positives and false negatives (i.e., the percent of ASD individuals that match the conjunctive clause). In this work, the CCEA was run five times using the training set to ensure a more thorough search of the fitness landscape.

### K-nearest Neighbors Algorithm and Leave-One-Out Cross Validation

In order to further validate the CCEA’s selection of features capable of discriminating between children with and without ASD, we built a separate KNN classification model and used leave-one-out cross validation on all 28 subjects. The KNN is a classification algorithm that assumes that things that exist in close proximity (i.e., nearer to each other) are more similar. In this study, each subject was classified into one of two output classes (i.e., ASD or NT) based on a plurality vote of its neighbors, with the subject being assigned to the class most similar to its k-nearest neighbors. When k = 1, then the subject is assigned to the class of a single nearest neighbor [65]. Leave-one-out cross validation is a special case of cross validation where the number of folds equals the number of subjects in the data set. Thus, the KNN algorithm is applied once for each subject, using all other instances as the training set and using the selected subject as a single-item test set [66]. After model validation, we trained three separate KNN classifiers using a balanced dataset (6 NT and 6 ASD subjects) and feature sets identified by the CCEA to classify the remaining 16 subjects. See Table 2.

**Table 2:** Subject Inclusion and Distribution.

| Models |  | Subjects | Include Subjects Used In CCEA?<br>N=21 (14 NT & 7 ASD) | Include Subjects from the Later Cohort That Was Not Used In CCEA?<br>N=7 (5 NT & 2 ASD) |
| --- | --- | --- | --- | --- |
| CCEA |  | NT = 14<br>ASD = 7 | All | None |
| KNN Validation Model |  | NT = 19<br>ASD = 9 | All | All |
| KNN Prediction Model | Training | NT = 6<br>ASD = 6 | NT = 6<br>ASD = 4 | NT= None<br>ASD = 2 |
|  | Testing | NT = 13<br>ASD = 3 | NT = 8<br>ASD = 3 | NT = 5<br>ASD = None |

## Results

### CCEA Feature Selection: 14 NT and 7 ASD

When using the CCEA for feature selection, 2438 CCs (i.e., models or sets of features) were generated ranging from first-order to fifth-order. The PPV of the 2438 models ranged from 46.47% to 100% and their class coverage ranged from 42.86% to 100%. Among these models, we looked for the most parsimonious (i.e., lowest order models) to draw meaningful conclusions and to avoid the overfitting that often occurs with higher-order models. As a result, we selected 8 second-order models (i.e., those having only two features) with the highest fitness (PMF) among the total 520 second-order models. These 8 “best performing” models each have 100% PPV and 100% class coverage, see Table 3. All of the features identified were brain anatomical features.

**Table 3.** Second-order CC model features and range of values.

| CC # | Feature One | Value | Feature Two | Value |
| --- | --- | --- | --- | --- |
| 113 | lh posteriorcingulate volume | [3500,4600] | lh rostralmiddlefrontal volume | [20000,25000] |
| 679 | lh posteriorcingulate volume | [3500,4600] | rh rostralmiddlefrontal volume | [21000,26000] |
| 1449 | lh posteriorcingulate volume | [3500,4600] | lh medialorbitofrontal thickness | [2.6,2.8] |
| 2199 | lh posteriorcingulate volume | [3500,4600] | rh rostralmiddlefrontal area | [6400,8500] |
| 887 | rh isthmuscingulate volume | [3300,4100] | rh posteriorcingulate volume | [4100,6200] |
| 1488 | rh isthmuscingulate volume | [3300,4100] | lh medialorbitofrontal thickness | [2.6,2.8] |
| 2200 | rh isthmuscingulate volume | [3300,4100] | rh rostralmiddlefrontal area | [6400,8500] |
| 2269 | rh isthmuscingulate volume | [3300,4100] | rh posteriorcingulate area | [1400,2900] |

Using CC 113 (Table 3) as an example, this second-order model can be interpreted as: any subject whose posterior cingulate gyrus volume was within the range of 3500 to 4600 *mm*^3^ AND left rostral middle frontal gyrus volume was within the range of 20,000 to 25,000 *mm*^3^ would be classified as having ASD. The volume of the left hemisphere posterior cingulate gyrus and the volume of the right hemisphere isthmus of the cingulate gyrus were the two features to appear most frequently (i.e., four times) across all second-order models, suggesting that the volume of cingulate gyrus is a potentially important biomarker for ASD. Figure 1 provides a 2D visualization for the range of feature values (numerical boundaries) associated with these models and the placement of each subject within this range.

**Fig 1.**
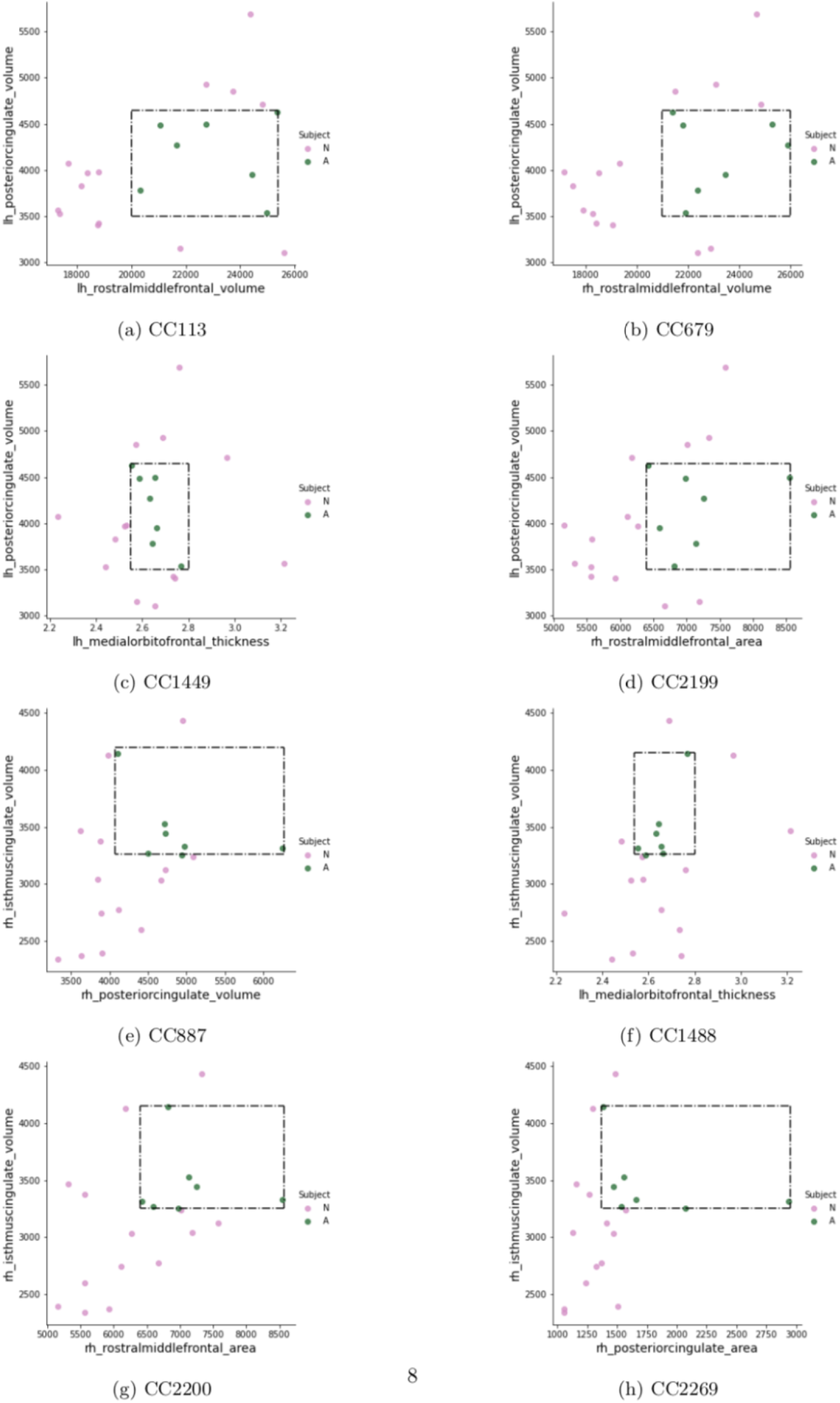
2D visualization of second-order CC models. Green dots represent ASD subjects and group together within the rectangle defining the range of values in Table 3.

Because of our desire to explore the predictive capability of the behavioral features, we expanded our analysis to include third-order models (i.e., model combinations with three features). There were 651 third-order models in total; while some consisted only of 310 anatomical brain features, others included two behavioral features plus one brain anatomical feature. We selected the 6 best-performing third-order models with the highest fitness (PMF); each had 100% PPV and 100% class coverage, see Table 4. Each of these third-order models contained two behavioral features and one brain anatomical feature.

**Table 4.** Third-order CC model features and range of values.

| CC # | Feature One | Value | Feature Two | Value | Feature Three | Value |
| --- | --- | --- | --- | --- | --- | --- |
| 46 | ToMTB total | [5,13] | ToMI early subscale mean | [12,18] | lh parsorbitalis meancurv | [0.17,0.2] |
| 191 | ToMTB total | [5,13] | ToMI early subscale mean | [12,18] | rh superiorparietal thickness | [2.4,2.7] |
| 264 | ToMTB total | [5,13] | ToMI early subscale mean | [12,18] | rh parsorbitalis meancurv | [0.18,0.2] |
| 317 | ToMTB total | [5,13] | ToMI early subscale mean | [12,18] | rh inferiortemporal meancurv | [0.15,0.18] |
| 1163 | ToMTB total | [5,13] | ToMI total composite mean | [9,18] | lh postcentral thickness | [2.2,2.5] |
| 1446 | ToMTB total | [5,13] | ToMI early subscale mean | [12,18] | lh medialorbitofrontal thickness | [2.6,2.8] |

Using CC 46 (Table 4) as an example, any subject who had a total score on ToMTB within the range of 5 to 13 AND an early subscale mean score on ToMI-2 within the range of 12 to 18 AND a mean curvature value of the left hemisphere pars orbitalis within the range of 0.17 to 2 would be classified as having ASD. The ToMTB total score feature occurred in all of our best-fit, third-order models; and the ToMI-2 early subscale mean score occurred in all but one (CC 1163) of the models, where the ToMI-2 total composite mean played a role. Such a finding further suggests that the ToMTB and ToMI-2 might be effective for ASD testing and diagnosis. See Figure 2.

**Fig 2.**
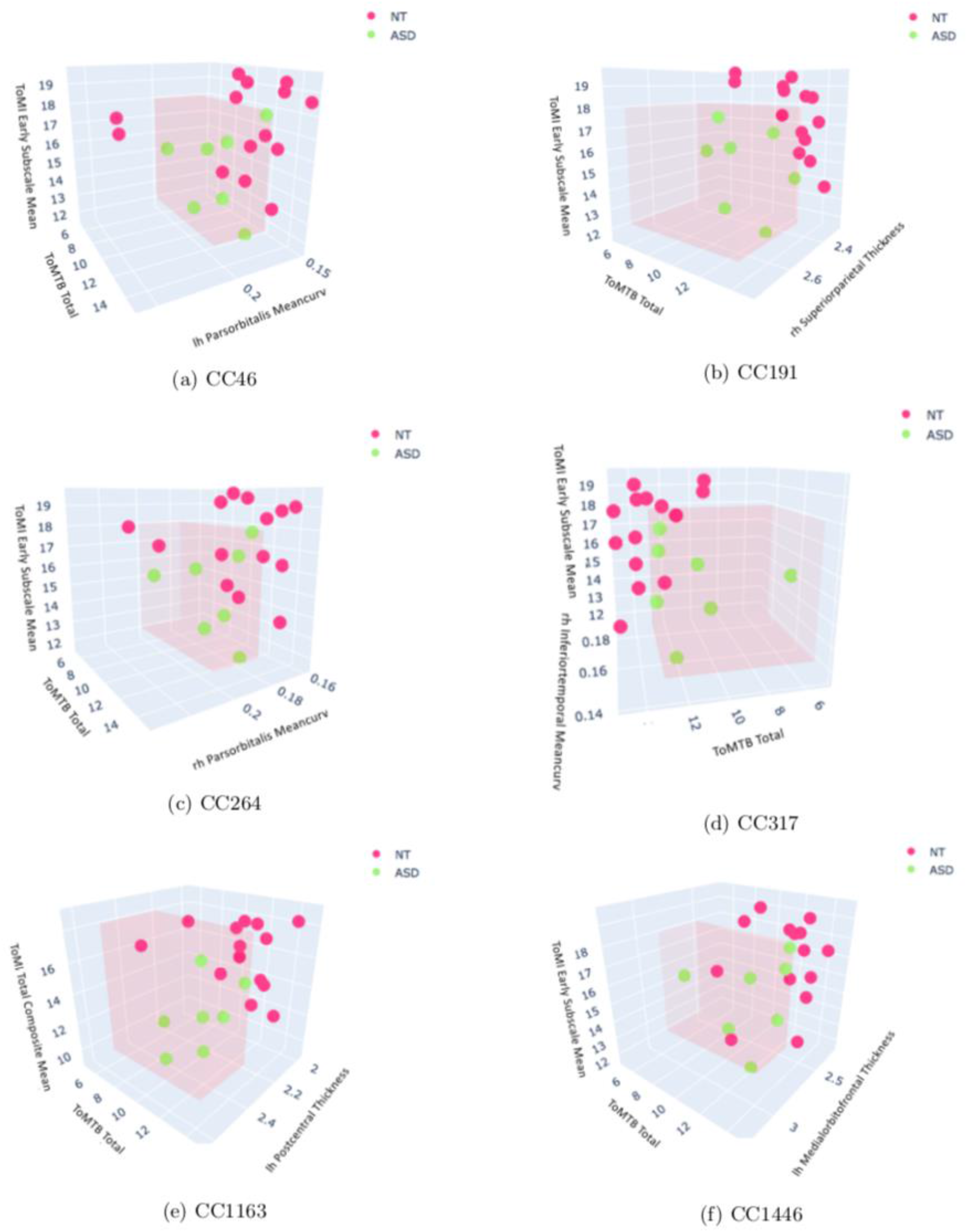
3D visualization of third-order CC models. Green dots represent ASD subjects and group together within the pink cube defining the range of values in Table 4.

### KNN Leave-One-Out Cross Validation

As mentioned earlier, a cohort of new subjects comprising 2 ASD and 5 NT children were enrolled at a later time. This later cohort was combined with the 21 subjects used in CCEA to cross-validate the KNN classifiers. Using the 8 unique features of the second-order model, the KNN (k=3) achieved 89.29% classification accuracy, where 17 of the 19 NT subjects and 8 of the 9 ASD were classified accurately; see Table 5, diagonals of the second-order confusion matrix.

**Table 5.**
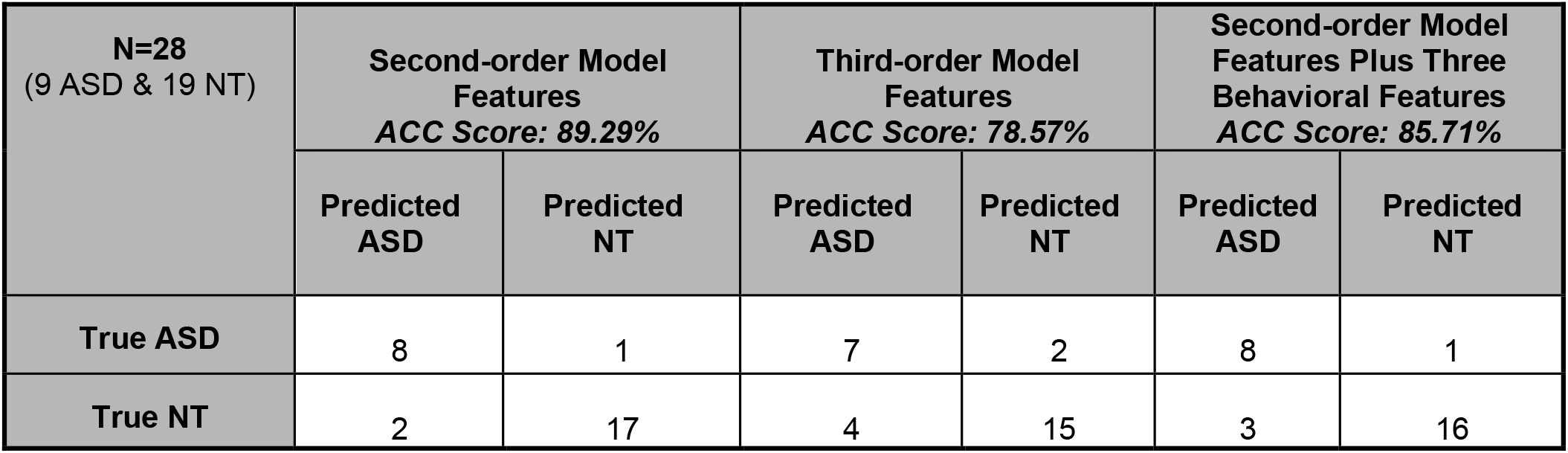
Cross Validation Confusion Matrices.

Using the 9 unique features of the third-order model, the KNN (k=7) validation accuracy fell to 78.57% compared to when using the 8 unique features from the second-order model, where 15 of the 19 NT and 7 of the 9 ASD subjects were classified accurately. See Table 5, diagonals of third-order confusion matrix.

Given the better ASD classification performance of the second-order neuroanatomical features, and that the behavioral measurements/features are relatively easier to collect among children with ASD, it was important to explore whether the ASD prediction results might be improved when the behavioral features were combined with the second-order features. As a result, we added the three behavioral features (i.e.,ToMTB total score, ToMI-2 total composite mean and ToMI-2 early subscale mean) from the third-order models to the 8 second-order brain anatomical features and cross-validated a new KNN (k=2) classifier. With the total of 11 unique features, a validation accuracy of 85.71% was achieved with 16 of the 19 NT subjects and 8 of the 9 ASD being classified accurately. See the confusion matrix of Table 5.

### Classification of ASD and NT subjects using the KNN model

To further examine whether the KNN classifier could discriminate subjects with ASD and NT, we developed three classification models – one using the 8 unique features from the second-order models, one using the 9 features from the third-order models, and one using the 11-feature model (i.e., eight second-order neuroanatomical features and three behavioral features). The best classification accuracy was achieved using a balanced training set that consisted of 6 NT and 6 ASD subjects, among which only 2 of the 6 ASD subjects were not part of the original CCEA feature selection. The remaining 16 subjects were used for testing (13 NT and 3 ASD).

The KNN (k=1) results for the second-order model features are shown in Table 6, columns 2 and 3; a classification accuracy of 87.5% was achieved, with all 3 of the ASD subjects and 11 of the 13 NT being classified accurately. Both of the misclassified NT subjects were part of the original CCEA feature selection.

**Table 6.** Classification Confusion Matrices.

| N=16<br>(3 ASD & 13 NT) | Second-order Model<br>Features<br>ACC Score: 87.5% |  | Third-order Model<br>Features<br>ACC Score: 81.25% |  | Second-order Model<br>Features Plus Three<br>Behavioral Features<br>ACC Score: 93.75% |  |
| --- | --- | --- | --- | --- | --- | --- |
|  | Predicted<br>ASD | Predicted<br>NT | Predicted<br>ASD | Predicted<br>NT | Predicted<br>ASD | Predicted<br>NT |
| True ASD | 3 | 0 | 3 | 0 | 3 | 0 |
| True NT | 2 | 11 | 3 | 10 | 1 | 12 |

The KNN (k=3) classification accuracy for the third-order model features was 81.25%, with all 3 of the ASD subjects and 10 of the 13 NT subjects correctly classified. Of the 3 misclassified NT subjects, 2 were included in the original CCEA feature selection. See Table 6, columns 4 and 5.

Lastly, the KNN (k=3) predictions for the combined 11-feature model (8 neuroanatomical features and three behavioral features) are shown in Table 6, columns 6 and 7; classification accuracy is 93.75%, with all 3 of the ASD subjects and 12 out of 13 NT subjects classified accurately. The one misclassified NT subject was not part of the original CCEA feature selection analysis.

## Discussion

This study used a new ML feature selection tool, the CCEA, to identify biomarkers and behavioral features capable of successfully discriminating between children (7 to 14 year of age) with and without ASD given a small dataset collected from a single research site. ML tools have long been applied to ASD research, but it remains a far-reaching goal to build a diagnostic system for ASD that incorporates both feature selection and prediction. Previous studies face the challenge of using datasets across different research sites for classification purposes, rather than identifying input variables most closely associated with ASD (i.e., feature selection) [17, 18, 32–35, 37–39, 67, 68]. Additionally, traditional ML algorithms do not work well with ASD datasets given the large amount of variance and the heterogeneous nature of the disorder [40, 41]. Meanwhile, it requires tremendous effort to include ASD individuals in a research study given the social, cognitive, and language challenges of this population. Thus, nearly all ASD datasets have a large number of features with relatively small sample sizes which, despite being unsuitable for many ML algorithms, often leads to overfitting and poor classification accuracy. However, the CCEA in this work was able to address such issues by efficiently exploring large search spaces for feature interactions associated with some nominal outcome (e.g., ASD or NT). It also adopted built-in tools to prevent overfitting to produce parsimonious models.

The present study demonstrated exceptionally good performance (i.e., 100% ASD PPV and 100% class coverage) of the features identified by the CCEA. The selected CCEA features from the parsimonious second and third order models included volume, area, cortical thickness, and mean curvature in specific regions around the cingulate cortex, frontal cortex, and temporal-parietal junction areas as biomarkers for ASD (e.g., the pericalcarine cortex, posterior cingulate cortex, isthmus of the cingulate gyrus, pars orbitalis, etc.). Such findings are consistent with previous literature suggesting that individuals with ASD have abnormalities in these brain regions [17, 21– 28, 69–72], which facilitates our identification of potential ASD biomarkers. Additionally, third-order models from the training set include measurements from the ToMI-2 and the ToMTB as important features [50, 53]. This provides evidence that outcome measures from ToMI-2 and ToMTB are able to distinguish ASD from NT which further validates the use of these tools in ASD assessments.

It is impressive that the KNN classifiers are able to achieve such high classification accuracy given our sample size, and validation of the discriminant features selected by the CCEA models. In particular, the KNN classifiers perform better using the second-order neuroanatomical features than the third-order feature models, which emphasizes the importance of focusing on parsimonious models selected by the CCEA. In addition, the KNN achieved the highest classification accuracy when adding the behavioral features from the third-order models to the neuroanatomical features from the second-order models. In most cases, neuroimaging measurements are conducted along with behavioral assessments. As the third-order models are included to explore the potential role that behavioral features might play along the side of neuroanatomical features, it is convincing to see the highest classification accuracy achieved when combining the behavioral features and the neuroanatomical features. These findings highlight the heterogeneous and multi-facet characteristics of ASD itself. Thus, although it is more difficult to implement MRI among children with ASD, such findings support the idea that neuroanatomical measurements increase confidence in diagnosis. It also suggests that a good ASD prediction model should consider including both behavioral and neuroanatomical features to establish brain-behavior connections to advance our understanding of ASD.

This study further demonstrates the robustness of the CCEA as a feature selection methodology. The accuracy of these features when used as input variables in the KNN classifier suggests their potential to help clinicians and researchers target specific domains in ToM in treating the social challenges most often seen in children with ASD. The implications of our findings for clinical researchers reinforce earlier findings regarding the brain-behavior connections for children with and without ASD related to ToM understanding [49, 51, 53, 73–75]. Knowing these connections may guide future researchers in the assessment of change following intervention at both a behavioral and neurobiological level. This may also lead to knowledge about which interventions may be most effective for children with specific neurobiological markers.

The present study has established important biomarker candidates of ASD. These biomarker candidates support previous research adopting traditional neuroimaging measurements identifying similar brain regions to explain the abnormalities in ASD [17, 21–28, 71]. Importantly, ML methodologies can perform as well as the traditional approaches in the field of neuroscience and specifically in our assessment of ASD in selecting neuroanatomical biomarkers. Although ML techniques have been adopted to help with diagnosis and treatment development in medicine [76– 79], the heterogeneity in ASD creates challenges. Typically, large, diverse, and comprehensive datasets are required to extract solid biomarkers, which can be time-consuming and may be less accurate with traditional approaches. Under such circumstances, ML techniques as described in this study can help advance the development of an automatic diagnostic and predictive system for ASD. The present study provides a new direction for adopting ML techniques in ASD research and other areas of medicine with similar heterogeneity in disease conditions.

## Data Availability

The data in this manuscript was collected by the research team under Dr. Patricia Prelock, access can be granted upon request.

## Acknowledgments

We thank Jay V. Gonyea, Administrative Director, and Scott Hipko, Senior Research Technologist, in the MRI Research Unit at the University of Vermont, for their support in acquiring the MRI scans. We thank Dr. Richard Watts, Ph.D., Director of the FAS Brain Imaging Center at Yale University, and Dr. Joseph Orr, Ph.D., Assistant Professor at Texas A&M University, for sharing their knowledge in MRI data pre-processing.

## Notes

### Competing Interest Statement

The authors have declared no competing interest.

### Funding Statement

This project was supported by a private donor committed to advancing research in autism spectrum disorder, as well as pilot MRI scan research funding from the MRI research unit at the University of Vermont.

### Author Declarations

The research is approved by the IRB office at the University of Vermont, protocol number is 19-0005.

### Summary of Updates

Added additional analysis

